# Utility of 3D printing in the surgical management of intra-articular distal humerus fractures: A protocol for Systematic Review and Meta-analysis

**DOI:** 10.1101/2022.01.25.22269836

**Authors:** Vishnu Baburaj, Sandeep Patel, Vishal Kumar, Mandeep Singh Dhillon

## Abstract

**Background:** The quality of reduction in distal humerus fractures plays a crucial role in obtaining good postoperative functional outcomes. The use of 3D printed models for preoperative planning could help improve reduction and hence potentially influencing outcomes after surgery.

**Objectives:** This systematic review aims to compare the outcome of surgeries done with the help of 3D printing assistance to those done with conventional techniques in distal humerus intraarticular fractures.

**Methods:** A systematic review will be conducted according to the PRISMA guidelines. Electronic databases of PubMed, Embase, Scopus and Ovid will be searched with a pre-defined search strategy. Bibliography of included articles will also be searched for more potential studies. Original research articles published since inception till the date of search in English language that compare the outcomes of 3D print-assisted surgery with those done without the help of 3D printing will be included. Review articles, conference abstracts and animal studies will be excluded. Outcome measures will be extracted from the selected studies and analysed with the help of appropriate software.

## 1. Background

Intraarticular distal humerus fractures are rare injuries that pose a challenge for the operating surgeon due to the complex anatomy. The preferred modality of treatment in young patients is open reduction and internal fixation (ORIF). The outcomes after ORIF are directly influenced by the quality of initial reduction, as is the case for most intraarticular fractures [1, 2]. 3D printed models could prove to be immensely helpful in the preoperative planning, by allowing the surgeon to physically feel and manipulate the various fracture fragments and simulate reduction. This could potentially improve the reduction obtained during the actual surgery thereby improving outcomes.

## 2. Need for review

There is limited literature on the use of 3D printed models in distal humerus fractures and no systematic reviews done till date. Hence this systematic review and meta-analysis aims to give a comprehensive overview of the application of 3D printing in the surgical management of distal humerus fractures.

## 3. Objectives

- To provide a detailed outline on the current applications of 3D printing in distal humerus fractures
- To compare outcomes of surgeries done with the help of 3D printed models to those done without using 3D printing

## 4. PICO framework for the study

a. Participants : Adult human subjects with a distal humerus fracture
b. Intervention : Surgery done with 3D printing assistance
c. Control : Surgeries done conventionally without using 3D printing
d. Outcome : Mean operating time, mean blood loss, radiation time, complications, functional outcomes and union rates.

## 5. Methods

This systematic review and meta-analysis will be conducted in accordance with the Preferred Reporting Items for Systematic Reviews and Meta-analysis (PRISMA) guidelines.

### a. Review Protocol

A protocol of the review will be prepared adhering to the PRISMA-P guidelines.

### b. Eligibility Criteria

Any original research on adult human subjects that compare surgeries done using 3D printed models for preoperative planning with those done without the help of 3D printing will be included. Studies in languages other than English, animal studies, review articles, conference abstracts will be excluded.

### c. Information Sources & Literature search

A literature search will be conducted on the following electronic databases of PubMed, Embase, Scopus and Ovid using the search string “(((3D print*) AND (distal OR intercondylar OR intraarticular)) AND (humerus)) AND (fracture)”. A manual reference search of the included articles will also be carried out for potentially eligible articles. Articles published from inception till date of search will be included.

### d. Study Selection

Two independent authors will screen the title and abstract of all articles obtained from the initial search. Full text of potentially eligible articles will be obtained and evaluated according to the inclusion and exclusion criteria. Any conflict of judgement with regard to inclusion or exclusion of any articles will be resolved by discussion between the authors.

### e. Data Collection & Data Items

Date will be extracted on specially designed data collection excel spreadsheets, and will be cross checked for accuracy. The data that will be collected will include:

- Name of first author and year of publication
- Study design
- Number of participants, their mean age and sex distribution
- Average duration of surgery
- Mean blood loss
- Rate of complications
- Postoperative range of motion
- Outcome at final follow up

### f. Outcome Measures

The following outcome measures will be evaluated:

- Mean operating time
- Mean blood loss
- Radiation exposure
- Quality of reduction
- Rate of complications
- Final functional outcome

### g. Data Analysis and Synthesis

Qualitative and qualitative synthesis will be carried out. Qualitative synthesis will be presented with the help of tables and figures depicting the main findings of the included studies. Data extraction will be carried out with the help of predesigned extraction forms. Quantitative analysis will be done to ascertain the pooled estimates of outcome parameters if it is reported by at least 3 studies. Heterogeneity will be reported using the *I*^*2*^ statistic. A fixed-effects model will be used in case heterogeneity is less than 50%, and a random-effects model if heterogeneity comes out to be greater than 50%, using 95% confidence intervals. Mean difference will be used for continuous variables and odds ratio will be used for dichotomous variables. Results will be depicted with the help of forest plots. Meta-analysis will be carried out using RevMan version 5.4.

### h. Risk of bias

Risk of bias assessment will be done using MINORS tool for observational studies and ROB 2.O tool for randomised controlled trials [3, 4].

## Data Availability

All data produced in the present work are contained in the manuscript

